# Characteristics of adolescents presenting with physical symptoms without organic etiology at the pediatric emergency department in Martinique

**DOI:** 10.1101/2025.06.12.25329452

**Authors:** Raïssa Antoy, Julien Denis

## Abstract

**Background:** Physical symptoms without an identified organic cause are frequently observed in adolescents. These symptoms are often linked to psychiatric comorbidities, posing diagnostic and management challenges for healthcare professionals.

**Objective:** To assess the proportion of pediatric emergency room visits for physical symptoms without organic etiology among adolescents with Patient Health Questionnaire-13 (PHQ-13) somatic symptoms. To assess the proportion of patients with somatic symptoms who received a psychosocial history, and describe the socio-demographic and clinical characteristics of these adolescents.

**Methods:** Quantitative cross-sectional descriptive study of adolescents consulting the pediatric emergency department in Martinique from February 1, 2023 to April 30, 2024. Inclusion criteria were patients aged 11 to 17 years consulting with a somatic symptom of the PHQ-13 questionnaire. Medical management in the emergency department was considered satisfactory if a psychosocial history based on the HEADSSS questionnaire was taken.

**Results:** 2398 emergency room visits were included. Analysis revealed that 42.2% of pediatric emergency consultations were related to symptoms without etiology. The majority of these consultations concerned girls (69.1 %, *p*≤ 0. 05). The HEADSSS questionnaire was used in only 6.4 % of cases (*p*≤ 0. 05). In our study, the most frequent physical symptoms without organic etiology were abdominal pain (35.7 %), chest pain (24.7 %) and malaise (24.7 %).

**Conclusion:** This study shows that consultations related to physical symptoms without an identified organic cause found in adolescents are frequent in pediatric emergency departments in Martinique. Their management appears to be sub-optimal, and prospects for improvement should be considered, such as setting up a management protocol, training doctors in this area and carrying out further prospective and interventional studies.

## INTRODUCTION

Physical symptoms without an identified organic etiology are common in the pediatric population, with a prevalence estimated between 10% and 30%, according to various studies (1,2). Adolescents frequently consult general practitioners and other medical specialists for physical symptoms with no identifiable organic etiology (3–5). Indeed, adolescents tend to present to somatic physicians rather than psychiatrists, as they often attribute their symptoms to a physical illness rather than a psychological one (6,7). The most commonly reported clinical manifestations involve gastrointestinal complaints (abdominal pain, transit disorders), cardiopulmonary symptoms (chest pain and malaise) or general bodily pain (arms, back and headaches) (6,7).

When these symptoms are not linked to a known medical condition, physicians often refer to them as “somatization” or “somatoform disorders” or “medically unexplained physical symptoms”. The use of these terms stems from the fact that modern medicine still operates within the Cartesian dualism of body and mind, in which illnesses are categorized as either “physical” or “psychological”. Polish psychiatrist Lipowski ZJ defined somatization as “a tendency to experience and communicate psychological distress in the form of physical symptoms” or “ to experience and express somatic distress in response to psychosocial stress” and “ to seek medical help” (8). Somatization is currently understood as the result of a complex interaction between the body and mind (9), which can lead to significant distress and functional problems in affected individuals (10). Therefore, it is essential to consider the impact of mental health in the expression of physical symptoms without an identified organic cause. This is especially critical given that the adolescent mental health has deteriorated in recent years, both in France and globally (11). Adolescence is a transitional period that inherently carries increased vulnerability to mental health issues.

When physical symptoms without an identified organic cause are associated with significant distress and impairment, they fall into the category of somatic symptom disorders and related disorders, as described in the DSM-5 (10). The editors of the DSM-5 have replaced the former term “somatoform disorders” with “somatic symptom disorders”, to reduce the stigma and acknowledge the validity of the symptoms experienced by patients (12). The diagnosis no longer relies solely on the absence of a clear medical explanation, but rather emphasizes the psychosocial distress caused by these symptoms – such as their impact on social and family life, academic performance, or the patient’s mood (12). When such symptoms persist for more than six months, the condition meets the criteria for somatic symptom disorders (SSD) in its full form (10,13). SSD is also common in adolescents (1,14). It represents a possible chronic evolution of unexplained physical symptoms and can have a considerable psychosocial impact on affected individuals.

Moreover, adolescents presenting with physical symptoms without an identified organic cause are at increased risk of developing other psychiatric comorbidities, such as anxiety disorders, depressive syndromes, or behavioral disorders (1). These comorbidities are known to be major risk factors for suicide (15). As a reminder, suicide is the second leading cause of death among 15-24 year-olds in France, following road traffic accidents (16). Therefore, it is essential to carry out early screening of adolescents with somatic symptoms without an identified organic etiology that have a psychosocial impact, in order to prevent progression toward other psychiatric comorbidities.

However, studies have shown that somatic physicians (general practitioners or other specialists) often experience difficulties in diagnosing and managing patients with physical symptoms without an identified organic cause (5,17). This may exacerbate the patient’s distress and increase the risk of developing other psychiatric comorbidities, particularly in adulthood (18,19). As a result patients frequently request further medical investigations from general practitioners in an attempt to find a physical explanation and receive a somatic-based treatment (20). Moreover, an English study found that even in the absence of explicit patient requests, general practitioners and other somatic physicians tend to focus on somatic management approaches (21). This suggests that they may resort to inappropriate or excessive testing and treatment.

In such circumstances, the management of physical symptoms in adolescents should not be limited to the search for somatic pathology, but should also include a psychosocial assessment. To facilitate this, there are validated screening tools that can be used in primary care settings or during brief consultations, such as in the pediatric emergency department. The Patient Health Questionnaire-13 (PHQ-13) is a tool designed to assess the severity of somatic symptoms (22). Another questionnaire, the HEADSSS, was specifically developed to identify mental health issues related to psychosocial factors in adolescents.

In Martinique, no recent studies have been conducted on the adolescent population consulting for physical symptoms without an organic etiology. Given the potential psychosocial impact and associated psychiatric co-morbidities, it seems essential to assess how such cases are managed in the pediatric emergency departments of the Maison de la Femme de la Mère et de l’Enfant. This is a necessary first step toward implementing early and appropriate care for these patients.

The primary objective of this study is to determine the proportion of consultation in the pediatric emergency department for physical symptoms without an identified organic etiology among adolescents presenting somatic symptoms as defined by the PHQ-13.

The secondary objectives are to evaluate the proportion of patients with somatic symptoms who received a psychosocial history, and to describe the socio-demographic, medical history, and clinical characteristics of these adolescents.

## MATERIALS AND METHODS

We conducted a descriptive cross-sectional quantitative study of patients consulting the pediatric emergency department at the Maison de la Femme de la Mère et de l’Enfant, within the University Hospital of Martinique (CHU de la Martinique).

### A. Study population

In this study, we included the medical records of patients aged 11 to 17 who consulted the pediatric emergency department between February 1, 2023, and April 30, 2024, for specific chief complaints. The 11-17 age range corresponds to the adolescent period in pediatrics (23).

Regarding the reason for consultation, we used the PHQ-13 as a reference. This pediatric self-questionnaire is derived from the PHQ-15 (24)(Appendix 1). The PHQ-15 is a validated tool used to assess the severity of common somatic symptoms and to screen for somatic symptom disorder (SSD)(22).

The 13 somatic symptoms included in the PHQ-13 questionnaire for adolescents are :

- Abdominal pain
- Back pain
- Limb pain
- Headache
- Chest pain
- Vertigo
- Malaise
- Palpitations
- Dyspnea
- Transit disorder
- Digest discomfort
- Asthenia
- Sleep disorder

Thus, our study included patients whose reason for consultation matched one of the symptoms listed in the PHQ-13. To identify these patients, we used regular expressions based on the presenting complaint as documented by the triage nurse.

We excluded patients who were not seen by a physician in the emergency department, those without a recorded reason for consultation documented by the triage nurse, and those whose medical records lacked physician notes.

### B. Data collection

Data collection was performed using the pediatric emergency management software Résurgences at the University Hospital of Martinique. Data extraction was conducted anonymously in csv format, followed by processing with custom Python scripts (notably using regular expressions) to automatically extract the variables of interest from text fields. Manual verification was then carried out through a Python-coded interface.

### C. Data analyzed

The variables of interest analyzed in this study are classified into the following categories :

#### 1. Socio-demographic charactéristics

We extracted data concerning gender, age, lifestyle (family structure, siblings, family atmosphere, recent moves and support persons), as well as the time and day of presentation to the emergency department.

#### 2. Medical characteristics including clinical and paraclinical management

We extracted data regarding the reason for consultation, patient history, presenting symptoms, abnormalities on clinical examination, biological tests, radiological examinations, ECGs, established diagnoses, and patient disposition (discharge, hospitalization, referral, no follow-up, etc.).

Patients were classified as either “with etiology” or “without etiology” based on whether an etiology was diagnosed during their visit. This classification was based on the diagnosis coded by the hospital practitioner : if the diagnosis contained any of the PHQ-13 keywords or referred to a presumed functional origin (e.g., “panic disorder”, “anxiety”, “stress”, “worry”, “anxious”, “panic”, “somatization”, “psychotic”, ‘depression’, “agitation”) the case was considered to lack an organic etiology.

#### 3. Psychosocial assessment of patients “without etiology”

To assess the quality of the psychosocial history of the adolescent presenting for physical symptoms “without etiology”, we relied on the items of the HEADSSS questionnaire. The HEADSSS questionnaire is used to assess mental health risk factors in the paediatric population (25). It explores psychosocial aspects of the patient’s life and helps determine whether additional psychiatric care is needed, particularly in a hospital setting. The questionnaire covers the patient’s family environment, educationnal status, nutrition, activities, sexual behavior, potential substance use, suicidal risk and safety concerns ( including risk-taking behavior, bullying, and abuse).

We searched the patients’ medical notes for mentions related to the items of the HEADSSS questionnaire, namely:

- Home : family structure; description of siblings; presence of tension or a negative family atmosphere (e.g, due to divorce, relocation, or other conflicts); existence of a trusted person for support; recent move; presence of a new household member or recent departure of another
- Education : current grade and academic track; academic performance quality; recent changes in grades; future plans; presence of friends at school; school absenteeism
- Eating : appetite; dysmorphophobia; recent weight changes; appetite quality (anorexia, selective eating, fasting periods)
- Activities : hobbies; participation in organized club such as sports, music, or arts
- Drugs : use of toxic substance
- Sexuality : relationship status; sexually active; sexual orientation; protected sexual relations; contraception methods; history of sexual violence
- Suicide : sadness; suicidal thoughts; history of suicide attempts; presence or history of self-harm
- Safety : victim of bullying (school or otherwise); victim of physical violence; risk-taking behaviour

We considered the HEADSSS questionnaire to be adequately completed if at least one element from four out of the eight items was documented in the medical record.

#### 4. Assessment of the rate of suicide attempts following an emergency visit with no identified etiology

To assess this rate, we examined whether each patient classified as “without etiology” between February 1, 2023, and April 30, 2024, had a subsequent emergency department visit for, or mentioning, a suicide attempt. We used a set of keywords (“suicidal attempt”, “intentional medication overdose”, ‘suicide’, “intoxication”) in the medical reports to pre-select the records, which were then manually reviewed. If a patient had multiple visits for suicide attempts, only the first one in chronological order was retained for analysis.

### D. Statistical analyses

The study population was described using counts and percentage for qualitative and categorical variables, and by the median along with the 25th and 75th percentiles for continuous quantitative variables.

Comparative analyses for p-value calculation were conducted using a Mann-Whitney test for continuous quantitative variables, and the chi-square test for binary variables. A p-value less than 0.05 was considered statistically significant.

Effect sizes were calculated as standardized mean difference for continuous quantitative variables, and using Mahalanobis distance for binary variables, with corresponding 95% confidence intervals.

All analyses and figure generation were performed using custom Python code (Python 3.9). P-values were computed using the scipy package, and effect sizes were estimated using the effectsize package (26).

### E. Ethical and legal aspects

The study was conducted in accordance with French and international best practice guidelines for epidemiological research. Ethical approval was obtained following submission of the study protocol to the Institutional Review Board (IRB) (number 2024/049). Data analysis was carried out in compliance with confidence regulations, in line with the reference methodology proposed by the French Data Protected Authority “Commission Nationale de l’Informatique et des Libertés - CNIL”.

## RESULTS

### A. Study population

Among the 30,927 visits recorded at the pediatric emergency department of the University Hospital of Martinique between February 1, 2023, and April 30, 2024, a total of 2,398 visits met the inclusion criteria. The inclusion process is summarized in Figure 1. A total of 1,561 patients had a single visit to the pediatric emergency department during the inclusion period.

**Figure 1:**
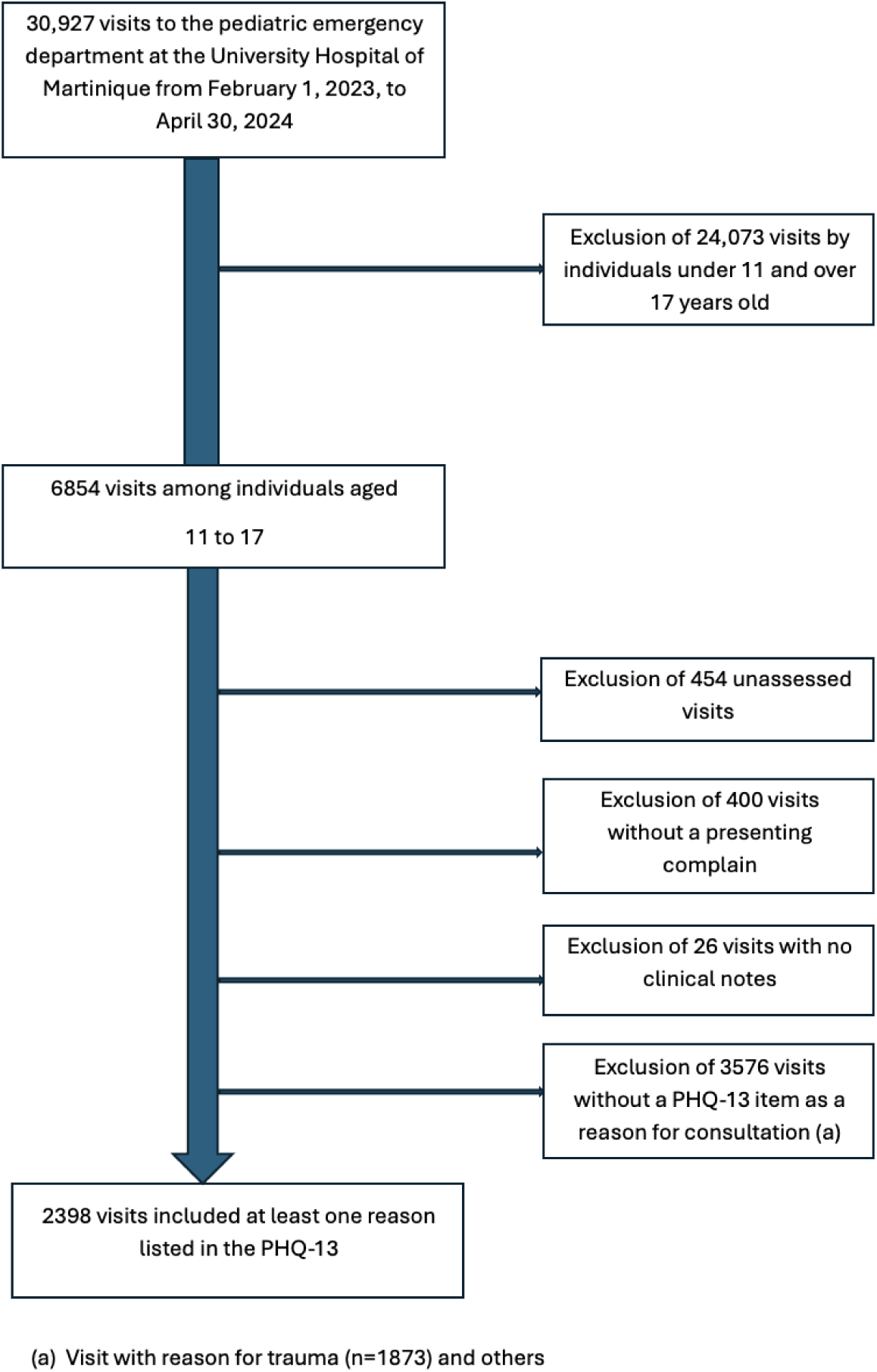
Study flow gram.

The socio-demographic characteristics of the included population are presented in Table 1.

**Table 1.** Socio-demographic characteristics.

|  | **Total sample size (%)** | **Without etiology (%)** | **With etiology (%)** | **p\*** | **Effect size [95.0 % CI]** |
| --- | --- | --- | --- | --- | --- |
| **Number of visits** | 2398 | 1011 (42.2) | 1387 (57.8) |  |  |
| **Median age** | 14.2 | 14.4 | 14.0 | <0.001 | 0.15 [0.07, 0.23] |
| **Age 11-14 years-old** | 1474 / 2398 (61.5) | 584 / 1011 (57.8) | 890 / 1387 (64.2) | 0.002 | 0.13 [0.05, 0.21] |
| **Age 15-17 years-old** | 924 / 2398 (38.5) | 427 / 1011 (42.2) | 497 / 1387 (35.8) | 0.002 | 0.13 [0.05, 0.21] |
| **Females** | 1444 / 2397 (60.2) | 699 / 1011 (69.1) | 746 / 1387 (53.8) | <0.001 | 0.32 [0.24, 0.4] |
| **Males** | 953 / 2397 (39.8) | 312 / 1011 (30.9) | 641 / 1387 (46.2) | <0.001 | 0.32 [0.24, 0.4] |

*p\* : With a p-value ≤ 0.05*

### B. Patients with physical symptoms without organic etiology

#### 1. Socio-demographic characteristics

According to Table 1, among the included adolescent population, 42.2% of the medical records corresponded to patients classified as “without etiology”. These consultations involved female patients in 69.1% of cases, with a median age of 14.4 years.

### 2. Psychosocial history and completion of HEADSSS questionnaire

Among visits with a PHQ-13 symptom as the reason for consultation, an assessment of at least four out of eight items from the HEADSSS questionnaire was completed in 3.5% of cases. The most frequently documented items in the records related to home, education and eating.

Females classified as “without etiology” were significantly more likely to have undergone HEADSSS evaluation (7.9%) compared to males (3.2%) (p=0.008) (Table 2). Detailed results by HEADSSS items for adolescents without etiology are presented in Appendix 2.

**Table 2.** HEADSSS questionnaire results by gender for adolescents with no etiology.

|  | Total sample size (%) | Without etiology females (%) | Without etiology males (%) | p* | Effect size [95.0 % CI] |
| --- | --- | --- | --- | --- | --- |
| **Number of visits** | 2398 | 699 (29.1) | 312 (13.0) |  |  |
| **HEADSSS Home** | 257 / 2398 (10.7) | 116 / 699 (16.6) | 32 / 312 (10.3) | 0.011 | 0.19 [0.06, 0.32] |
| **HEADSSS Education** | 225 / 2398 (9.4) | 103 / 699 (14.7) | 32 / 312 (10.3) | 0.067 | 0.14 [0.01, 0.27] |
| **HEADSSS Eating** | 785 / 2398 (32.7) | 187 / 699 (26.8) | 94 / 312 (30.1) | 0.302 | 0.07 [-0.06, 0.2] |
| **HEADSSS Activities** | 117 / 2398 (4.9) | 52 / 699 (7.4) | 33 / 312 (10.6) | 0.124 | 0.11 [-0.02, 0.24] |
| **HEADSSS Drugs** | 121 / 2398 (5.0) | 57 / 699 (8.2) | 22 / 312 (7.1) | 0.633 | 0.04 [-0.09, 0.17] |
| **HEADSSS Sexuality** | 176 / 2398 (7.3) | 106 / 699 (15.2) | 6 / 312 (1.9) | < 0.001 | 0.49 [0.35, 0.63] |
| **HEADSSS Suicide** | 170 / 2398 (7.1) | 102 / 699 (14.6) | 27 / 312 (8.7) | 0.012 | 0.19 [0.06, 0.32] |
| **HEADSSS Safety** | 97 / 2398 (4.0) | 60 / 699 (8.6) | 14 / 312 (4.5) | 0.029 | 0.17 [0.04, 0.3] |
| **HEADSSS ≥ 4 items /8** | 83 / 2398 (3.5) | 55 / 699 (7.9) | 10 / 312 (3.2) | 0.008 | 0.2 [0.07, 0.33] |

*p\* : With a p-value ≤ 0.05 ; ≥4 items /8 : if at least one element from four out of the eight items is documented, the HEADSSS assessment is considered to have been completed*

#### 3. Clinical characteristics

The most frequent reason for consultation at the triage nurse among patients classified as “without etiology” was abdominal pain (35.7%), followed by chest pain and malaise, each accounting for 24.7% of cases (Table 3).

**Table 3.** Results concerning the reason for consultation in the study population.

|  | **Total**<br>**sample size**<br>**(%)** | **Without**<br>**etiology**<br>**(%)** | **With**<br>**etiology**<br>**(%)** | **p\*** | **Effect size**<br>**[95.0 % CI]** |
| --- | --- | --- | --- | --- | --- |
| **Abdominal**<br>**pain** | 741 / 2398<br>(30.9) | 361 / 1011<br>(35.7) | 380 / 1387<br>(27.4) | <0.001 | 0.18<br>[0.1, 0.26] |
| **Chest pain** | 383 / 2398<br>(16.0) | 250 / 1011<br>(24.7) | 133 / 1387<br>(9.6) | <0.001 | 0.41<br>[0.33, 0.49] |
| **Headaches** | 482 / 2398<br>(20.1) | 130 / 1011<br>(12.9) | 352 / 1387<br>(25.4) | <0.001 | 0.32<br>[0.24, 0.4] |
| **Malaise** | 358 / 2398<br>(14.9) | 250 / 1011<br>(24.7) | 108 / 1387<br>(7.8) | <0.001 | 0.47<br>[0.39, 0.55] |
| **Back pain** | 160 / 2398<br>(6.7) | 57 / 1011<br>(5.6) | 103 / 1387<br>(7.4) | 0.099 | 0.07<br>[-0.01,0.15] |
| **Limb pain** | 263 / 2398<br>(11.0) | 60 / 1011<br>(5.9) | 203 / 1387<br>(14.6) | <0.001 | 0.29<br>[0.21, 0.37] |
| **Vertigo** | 128 / 2398<br>(5.3) | 43 / 1011<br>(4.3) | 85 / 1387<br>(6.1) | 0.054 | 0.08<br>[-0.0, 0.16] |
| **Palpitations** | 49 / 2398<br>(2.0) | 32 / 1011<br>(3.2) | 17 / 1387<br>(1.2) | 0.002 | 0.13<br>[0.05, 0.21] |
| **Dyspnea** | 490 / 2398<br>(20.4) | 105 / 1011<br>(10.4) | 385 / 1387<br>(27.8) | <0.001 | 0.45<br>[0.37, 0.53] |
| **Transit**<br>**disorders** | 150 / 2398<br>(6.3) | 50 / 1011<br>(4.9) | 100 / 1387<br>(7.2) | 0.029 | 0.09<br>[0.01, 0.17] |
| **Digestive**<br>**discomfort** | 104 / 2398<br>(4.3) | 46 / 1011<br>(4.5) | 58 / 1387<br>(4.2) | 0.737 | 0.02<br>[-0.06, 0.1] |
| **Asthenia** | 101 / 2398<br>(4.2) | 20 / 1011<br>(2.0) | 81 / 1387<br>(5.8) | <0.001 | 0.2<br>[0.12, 0.28] |

*p\* : With a p-value ≤ 0.05. No patient was recorded with sleep disorder.*

We aimed to compare the proportions of PHQ-13 symptoms reported during the anamnesis in consultations classified as with and without etiology, and to identify the most frequent symptoms among patients “without etiology”. Among females, sleep disorders, palpitations and chest pain were the most prevalent symptoms, with proportions 73%, 70%, and 70%, respectively (figure 2). Among males, palpitations, chest pain, and malaise were the most commonly reported symptoms, with proportions of 61%, 58%, and 53%, respectively (Figure 2).

**Figure 2:**
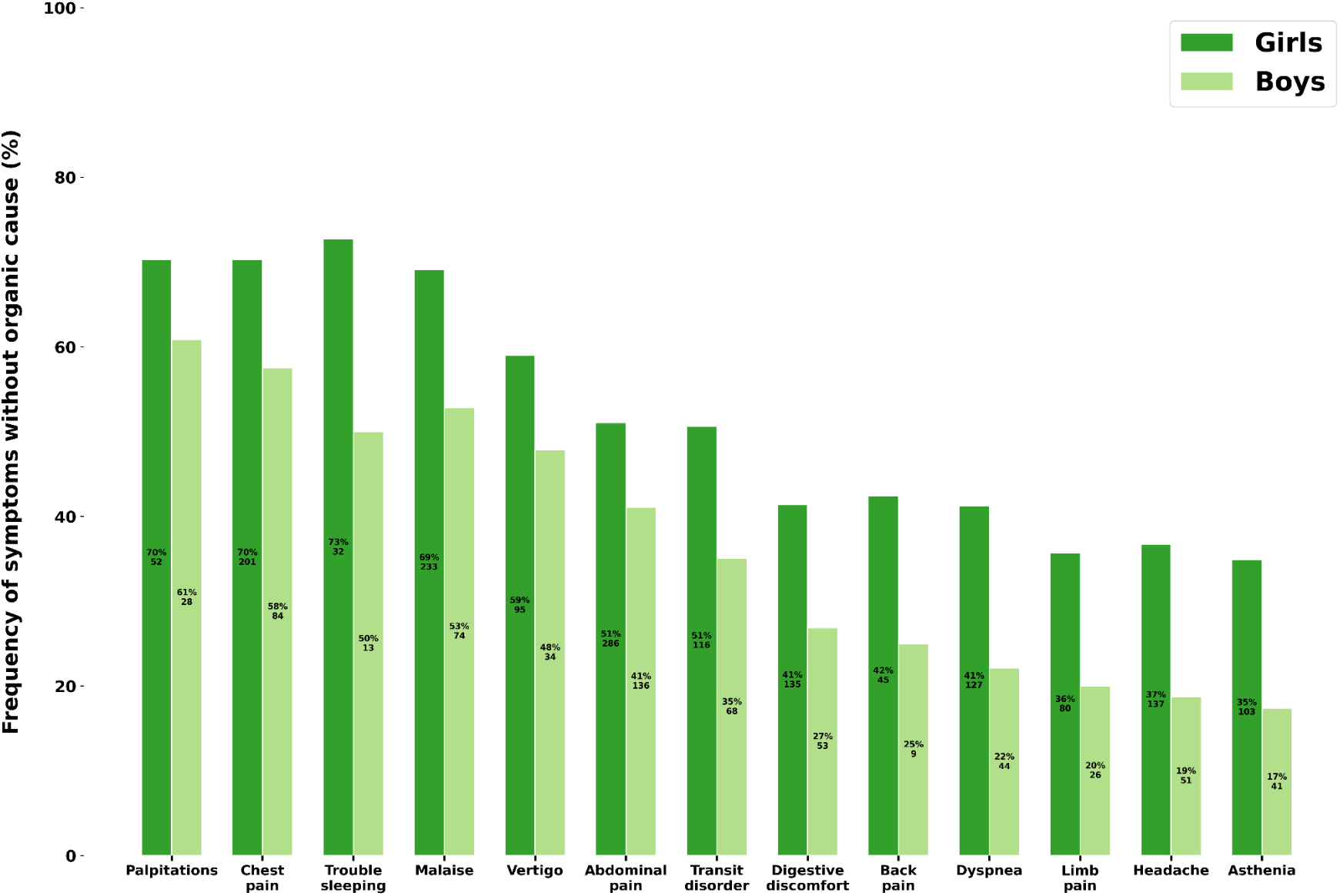
Frequency of symptoms without organic etiology (%) X-axis: symptoms reported during anamnesis (sorted in descending order) Y-axis: Proportion of cases without identified organic etiology for each symptom. For example, palpitations were reported in 74 consultations of female patients, among which 52 consultations (70%) had no identified organic etiology

#### 4. Complementary examinations

The results of complementary examinations performed in the paediatric emergency department are presented in Table 4. Biological tests were conducted in 40.6% of cases among patients classified as “without etiology”. Urine dipstick and electrocardiograms (ECG) were performed in 40.4% and 43.4% of these patients, respectively. Radiographs were obtained in 12.5% of cases. Other imaging examinations, such as ultrasound, CT-scan and magnetic resonance imaging (MRI), were carried out in 17.7% of these patients.

**Table 4.** Results of complementary examinations.

|  | **Total sample size (%)** | **Without etiology (%)** | **With etiology (%)** | **p\*** | **Effect size [95.0 % CI]** |
| --- | --- | --- | --- | --- | --- |
| **Rapid CRP test done** | 133 / 1404 (9.5) | 53 / 554 (9.6) | 80 / 850 (9.4) | 0.997 | 0.01<br>[-0.1, 0.12] |
| **Blood gas analysis done** | 253 / 1512 (16.7) | 82 / 583 (14.1) | 171 / 929 (18.4) | 0.033 | 0.12<br>[0.02, 0.22] |
| **Urine dipstick done** | 723 / 1980 (36.5) | 339 / 839 (40.4) | 384 / 1141 (33.7) | 0.002 | 0.14<br>[0.05, 0.23] |
| **Nasal swab done** | 44 / 1311 (3.4) | 1 / 502 (0.2) | 43 / 809 (5.3) | < 0.001 | 0.32<br>[0.21, 0.43] |
| **Biological tests done** | 869 / 2114 (41.1) | 342 / 842 (40.6) | 527 / 1272 (41.4) | 0.743 | 0.02<br>[-0.07, 0.11] |
| **Troponine test done** | 331 / 2398 (13.8) | 158 / 1011 (15.6) | 173 / 1387 (12.5) | 0.031 | 0.09<br>[0.01, 0.17] |
| **Dengue positive** | 225 / 2398 (9.4) | 0 / 1011 (0) | 225 / 1387 (16.2) |  |  |
| **ECG done** | 669 / 2398 (27.9) | 439 / 1011 (43.4) | 230 / 1387 (16.6) | < 0.001 | 0.61<br>[0.53, 0.69] |
| **Abnormal ECG** | 35 / 513 (6.8) | 13 / 345 (3.8) | 22 / 168 (13.1) | < 0.001 | 0.34<br>[0.15, 0.53] |
| **Radiographs done** | 259 / 2398 (10.8) | 126 / 1011 (12.5) | 133 / 1387 (9.6) | 0.030 | 0.09<br>[0.01, 0.17] |
| **Abnormal radiographs** | 32 / 259 (12.4) | 5 / 126 (4.0) | 27 / 133 (20.3) | < 0.001 | 0.52<br>[0.27, 0.77] |
| **Échography/ CT Scan/MRI done** | 382 / 2398 (15.9) | 179 / 1011 (17.7) | 203 / 1387 (14.6) | 0.05 | 0.08<br>[-0.0, 0.16] |
| **Abnormal echography/ CT-Scan/MRI** | 70 / 278 (25.2) | 7 / 129 (5.4) | 63 / 149 (42.3) | < 0.001 | 0.96<br>[0.71, 1.21] |
| **EEG done** | 65 / 2394 (2.7) | 44 / 1009 (4.4) | 21 / 1385 (1.5) | < 0.001 | 0.17<br>[0.09, 0.25] |

*p\* : With a p-value $\leq 0.05$ ; CRP : C-reactive Protein ; ECG : electrocardiogram ; MRI : magnetic resonance imaging ; EEG : electroencephalogram*

#### 5. Assessment of the rate of suicide attempts following emergency visits classified as “without etiolgy”

During the data collection period, thirteen patients presented to the emergency department for a suicide attempt., with a total of 29 prior visits classified as “ without etiology” before the suicide attempt. All of these patients were female, representing 1.51% of the 859 patients who had at least one visit without identified etiology.

The most frequent reasons for their visits were chest pain, headache and dyspnea (Figure 3).

**Figure 3:**
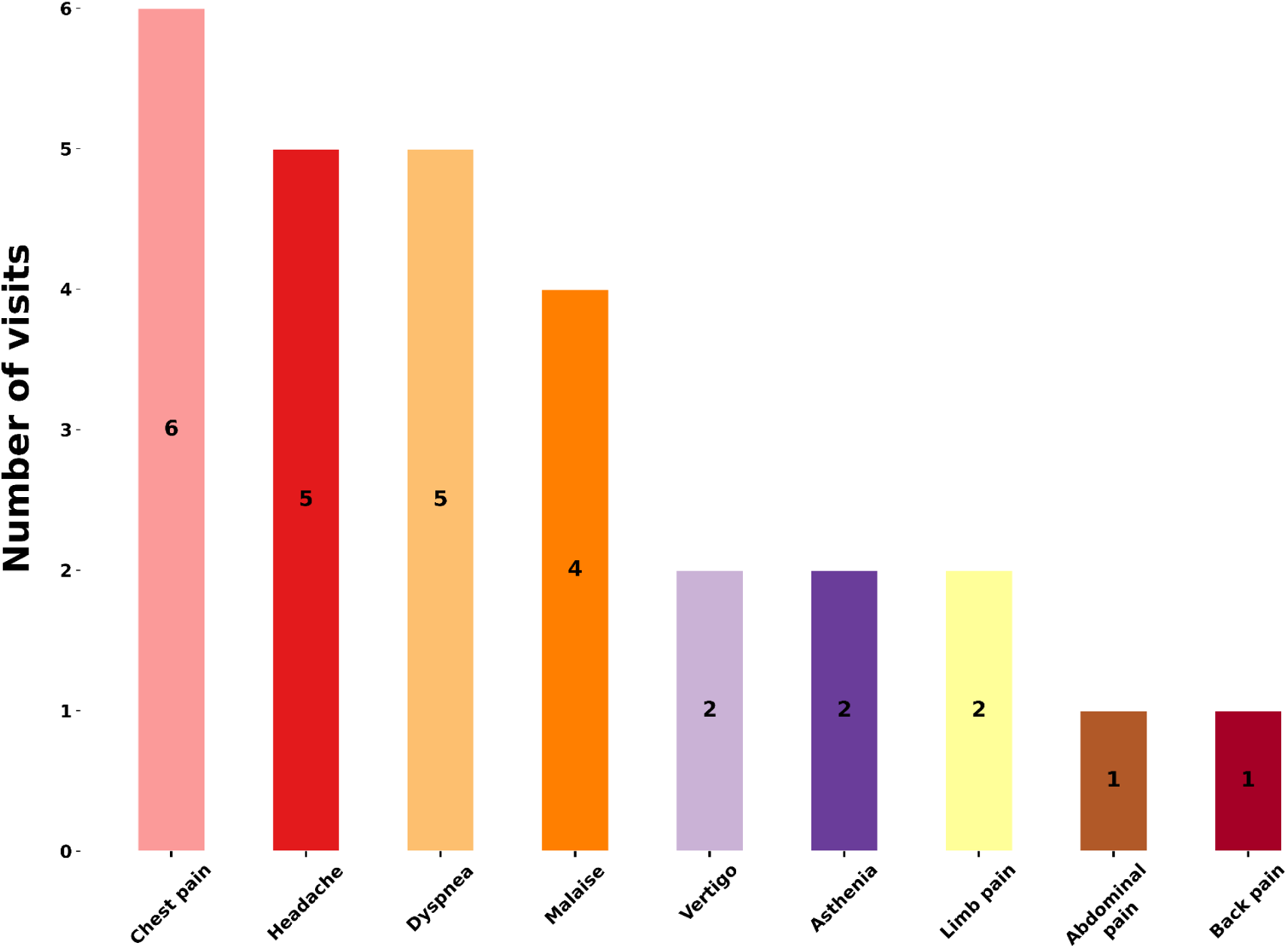
Number of visits by presenting complaints among patients prior to a suicide attempt.

During these visits, none of these patients had undergone an adequate evaluation using the HEADSSS questionnaire (defined as documentation of at least one element in at least four out of eight items).

### C. Differences between patients without and with identified etiology

#### 1. Socio-demographic characteristics

According to Table 1, a significantly higher proportion of females consulted for symptoms classified as “without etiology” (69.1%) compared to those “with etiology”. Males were significantly more likely to consult for complaints associated with an organic etiology (46.2%). Adolescents classified as “without etiology” presented significantly more often during school days, with a proportion of 58.5% (591 out of 1011), compared to 51.6% (715 out of 1387) among “with etiology”, (p < 0.001, effect size 0.14 [0.06 - 0.22]. Similarly, adolescents “without etiology” consulted significantly more frequently during school hours, with a proportion of 26.8% (271 out of 1011), versus 20.8% (289 out of 1387) for those “with etiology” (p < 0.001, effect size 0.14 [0.06 - 0.22].

#### 2. Psychosocial history and completion of the HEADSSS questionnaire

Adolescents classified as “without etiology” were significantly more frequently assessed using the HEADSSS questionnaire, with a proportion of 6.4% (65 out of 1011), compared to 1.3% (18 out of 1387) among adolescents “with etiology” (p < 0.05, effect size 0.27 [0.19 - 0.35]). Detailed results by HEADSSS questionnaire items for the study population are provided in Appendix 3.

#### 3. Clinical characteristics

According to Table 3, the most frequent reason for consultation among patients “with etiology” was dyspnea (27.8%), followed by abdominal pain (27.4%) and headache (25.4%). Headache and dyspnea were significantly more common among patients “with etiology”, whereas abdominal pain, chest pain, and malaise were significantly more frequent among patients “without etiology” (Table 3).

In anamnesis, girls and boys classified as “without etiology” most frequently presented symptoms such as abdominal pain, malaise and chest pain. Girls and boys classified as “with etiology” presented more commonly abdominal pain, headaches, and asthenia (Figure 4) (Appendix 4).

**Figure 4:**
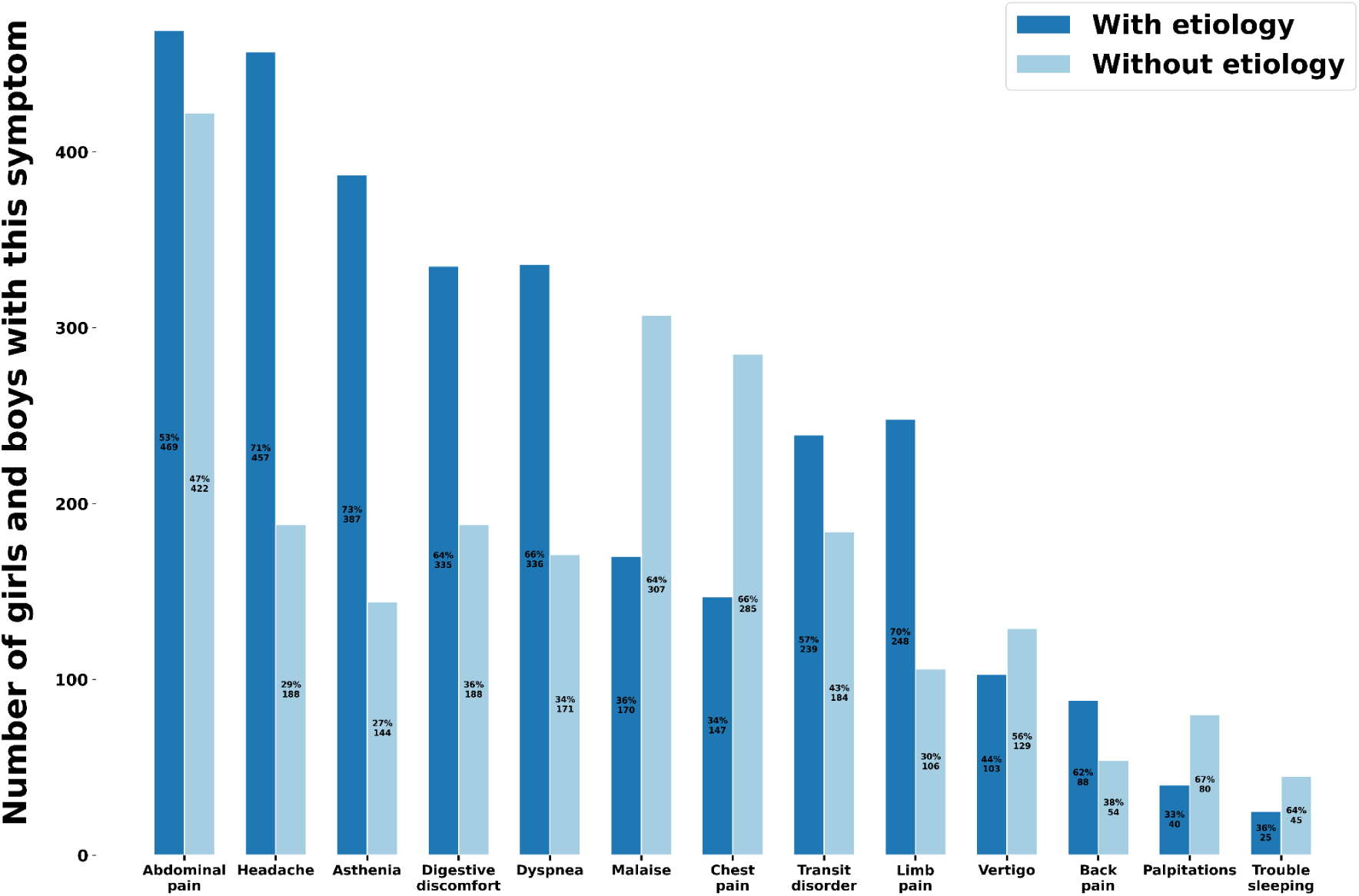
Number of adolescents per symptom with or without organic etiology. X-axis: symptoms found in the history (sorted in descending order) Y-axis: number of adolescent visits (ranked by frequency of patient presentations)

#### 4. Complementary examinations

According to Table 4, a significantly higher proportion of ECGs (43.4%), radiographs(12.5%) and other types of imaging (ultrasound, CT and MRI) (17.7%) were carried out in patients without an identified organic etiology. However, these examinations were significantly more likely to yield abnormal results in patients with a known etiology. There was no significant difference in the performance of blood tests between patients with and without an organic etiology.

## DISCUSSION

To our knowledge, this is the first epidemiological study conducted in Martinique describing the adolescent population presenting with physical symptoms without any organic etiology found. Our sample was representative, with 2,398 passages meeting the inclusion criteria among the adolescent population consulting at the pediatric emergency department of the MFME.

### Main objective

Our study revealed that 42.2% of consultations for somatic symptoms among adolescents had no identified organic etiology in the pediatric emergency department during the study period. These somatic symptoms without organic cause predominantly concerned females (69.1%), consistent with several studies (27–29). Female gender is frequently identified as a risk factor for the development of such symptoms (30).

### Psychosocial assessment and psychiatric comorbidities

Regarding the psychosocial evaluation related to mental health, our study shows that it was conducted in only 3.5% of all included visits and in 6.4% of cases among patients “without etiology”. The most frequently documented items in the medical records concerned the home environment, education, and nutrition. An interventional study conducted in an emergency department in England demonstrated that before implementing specific training on the HEADSSS, only 3% of adolescents received a psychosocial assessment, mainly focusing on home and education (31). After introducing a standardized HEADSSS form and a dedicated training session, the rate of psychosocial assessment increased to 35%. This study highlights the importance of developing similar initiatives to enhance psychosocial assessment of adolescents in pediatric emergency departments in Martinique.

Furthermore, it is possible that the psychosocial history was taken by the emergency physician but not thoroughly documented in the medical record. Additionally, classification bias may exist since we arbitrarily considered the HEADSSS questionnaire to be adequately completed if at least one item in four out of the eight domains was recorded.

During our study, we found that 1.51% of patients who consulted for symptoms without an identified organic etiology subsequently presented to the emergency department for a suicide attempt, and all were female. During their initial visits with unexplained somatic symptoms, no appropriate psychosocial assessment had been conducted by the physicians. This lack of assessment may reflect a clinical approach focuses primarily on somatic complaints, potentially resulting in a missed opportunity for early detection of suicide risk–especially when such risk manifests through physical sumptoms (32,33). This is particularly concerning given that the 2025 report from the French National Suicide Observatory, published by DREES, highlights a 40% increase in suicide rates among individuals under 25 since 2020, along with a rise in hospitalizations for self-inflicted injuries among teenage girls and young women (34).

Moreover, in our study, the rate of suicide attempts following emergency visits for symptoms without organic etiology was likely underestimated. We only searched for passages for suicide-related visits within the inclusion period and not beyond it. Additionally, not all suicide attempts result in emergency department consultations, which could further contribute to the underestimation of the phenomenon.

In our analysis, adolescents without an identified organic etiology were significantly more likely to present during school days (58.5%) and school hours (26.8%), with a *p* ≤ 0. 05. These findings suggest a possible link between school avoidance and the expression of somatic symptoms without an organic cause. A systematic review published in 2021 found a significant presence of somatic symptoms among students with school absenteeism (35). Furthermore, school bullying–recognized as a crime in France–has been identified as a potential factor to physical symptoms in pediatrics, as shown in a 2021 study (36,37). It therefore appears essential to investigate school-related factors when evaluating adolescents presenting with such symptoms.

### Clinical characteristics

In our study, the most frequently reported physical symptoms among adolescents without an identified organic etiology during their emergency visit were abdominal pain (35.7%), chest pain (24.7%) and malaise (24.7%). According to the literature, the most common symptoms in adolescents are headache, abdominal pain and limb pain (29,38,39).

In our study population, abdominal pain remained one of the most frequently reported symptoms. As a reminder, this is a common complaint among school-aged children and is often associated with psychological comorbidities (40). Therefore, consultations related to this symptom in adolescents should be considered as potential indicators of underlying psychosocial issues, warranting early and appropriate management. Conversely, our results showed that headaches were significantly more frequent in patients with an identified organic etiology (*p* ≤ 0. 05).

However, it is important to note that Martinique experienced a dengue epidemic in 2024. Since our study analyzed data from February 1, 2023, to April 30, 2024, the findings may have been influenced by the concurrent dengue epidemic. Indeed, dengue shares several symptoms with those assessed in the PHQ-13, such as headaches, abdominal pain, and malaise. We did not control for these potential confounding factors through multivariate analysis.

### Complementary examinations

Our study shows that, in the pediatric emergencies of the MFME, physicians more frequently prescribe complementary examinations–such as ECG (43.4%) , radiographs (12.5%,) or other type of imaging (ultrasound, CT and MRI) (17.7%) –for patients without an identified organic etiology, with *p*≤ 0. 05. However, these examinations revealed significantly fewer abnormalities compared to those performed in patients with an identified etiology. Paradoxically, it is important to note that psychosocial assessment using the HEADSSS questionnaire was rarely conducted in this group. This finding suggests a tendency toward overprescription of diagnostic tests at the expense of a potentially more appropriate psychosocial evaluation.

Regarding the performance of biological tests, our study did not find a significant difference between patients with and without an identified etiology. This may be related to the dengue epidemic that coincided with our study period. Indeed, it is possible that some of these tests were prompted by the suspicion of dengue, particularly in patients presenting with abdominal pain, headaches, with or without fever–symptoms that overlap with those included in the PHQ-13. We did not control for these confounding factors through multivariate analysis.

### Limitations of the study

Some limitations have already been discussed above, particularly those related to the specific challenges encountered during the study. We will now address the broader limitations of the study.

Our study presents several limitations, notably the presence of selection and information biases due to potentially incomplete, imprecise, or biased data. Medical records may contain errors, be poorly documented, or lack essential details. Additionally, a subjective bias may be present due to the selection of the chief complaint by the triage nurse. The coding of discharge diagnoses from the emergency department may also be subjective and lack accuracy.

Since our study is cross-sectional, we do not have access to follow-up data whether inpatient or outpatient. We have no information on whether an organic etiology may have been identified later on. The lack of follow-up prevents us from identifying hidden or intermediate factors that could explain the observed results.

There is also a confounding bias due to the concurrent dengue epidemic. Given the design of our study, no causal relationship can therefore be established.

Furthermore, as our study was limited to a pediatric emergency department in Martinique, its external validity is restricted. Larger, multicenter studies would be necessary to validate and generalize these findings.

### Reflection axis

Although somatic symptoms affect a considerable proportion of adolescents, there is no consensus management approach in the literature. In any case, the systematic consideration of the psychosocial dimension in these adolescents is essential, given the associated psychiatric comorbidities. The use of questionnaires such as the PHQ-13 and the HEADSSS appears well-suited for the early detection of these symptoms before any significant psychosocial impact develops. Indeed, these tools have the advantage of targeting the specific needs of the patient and guiding appropriate care. A shortened version of the HEADSSS, specifically designed for use in emergency departments, has been developed under the name HEADS-ED score (Appendix 5). A score equal to or greater than eight and/or a suicide score of two increases sixfold the likelihood of requiring hospital care (41).

Furthermore, an Australian scientific article (7), published in 2018, recommends documenting in the patient’s medical record the suspicion of a diagnosis of physical symptoms without an identified etiology, when associated with psychosocial distress. This approach aims to reduce the overprescription of inappropriate investigations. In severe cases, the article also suggests a short hospital stay,, which may be necessary to rule out other possible psychiatric comorbidities and to initiate multidisciplinary support (pediatrician, psychiatrist, psychologist, general practitioner).

Similarly, it is also essential to strengthen the knowledge of somatic-care physicians (general practitioners or other specialists), as well as to improve the training of medical students in the management of these patients.

Finally, a prospective descriptive study–or even an interventional one–integrating appropriate screening tools for adolescents consulting for physical symptoms without an identified etiology in Martinique, could help better define their needs and improve their care.

## CONCLUSION

Our study highlights the significant proportion of consultations for physical symptoms without identified organic etiology among adolescents attending the pediatric emergency department of the MFME at the University Hospital of Martinique. These patients present with specific socio-demographic and clinical characteristics. However, their management within the MFME pediatric emergency department does not appear to be optimal. Given the risk of psychosocial impact and psychiatric comorbidities in these populations, improvement strategies should be considered, such as the implementation of a standardized care protocol and enhanced physician training on this issue. The management of adolescents presenting with physical symptoms without an identified organic cause, though complex, must follow a bio-psycho-social model and involve a multidisciplinary approach.

## Data Availability

All data produced in this study are available on reasonable request from the authors.

## Author contributions

**Raïssa Antoy**: Writing, review, editing, resources and visualization. **Julien Denis**: Methodology, software, formal analysis, validation, review and editing.

## APPENDIXES

## Appendix 1: PHQ-13: assessment of somatic symptom severity (24)

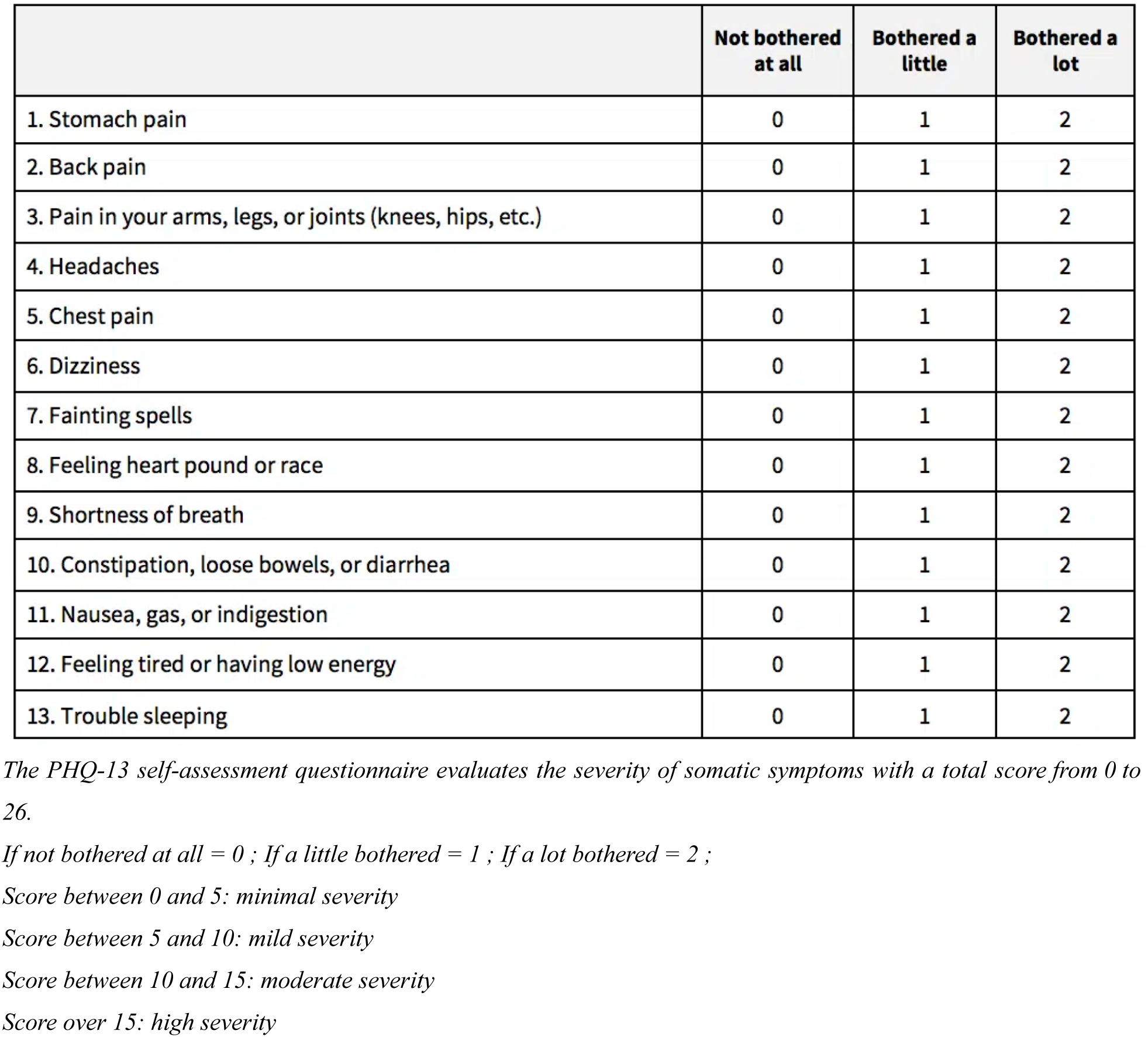

|  | Not bothered<br>at all | Bothered a<br>little | Bothered a<br>lot |
| --- | --- | --- | --- |
| 1. Stomach pain | 0 | 1 | 2 |
| 2. Back pain | 0 | 1 | 2 |
| 3. Pain in your arms, legs, or joints (knees, hips, etc.) | 0 | 1 | 2 |
| 4. Headaches | 0 | 1 | 2 |
| 5. Chest pain | 0 | 1 | 2 |
| 6. Dizziness | 0 | 1 | 2 |
| 7. Fainting spells | 0 | 1 | 2 |
| 8. Feeling heart pound or race | 0 | 1 | 2 |
| 9. Shortness of breath | 0 | 1 | 2 |
| 10. Constipation, loose bowels, or diarrhea | 0 | 1 | 2 |
| 11. Nausea, gas, or indigestion | 0 | 1 | 2 |
| 12. Feeling tired or having low energy | 0 | 1 | 2 |
| 13. Trouble sleeping | 0 | 1 | 2 |

*The PHQ-13 self-assessment questionnaire evaluates the severity of somatic symptoms with a total score from 0 to 26.*
*If not bothered at all = 0 ; If a little bothered = 1 ; If a lot bothered = 2 ;*
*Score between 0 and 5: minimal severity*
*Score between 5 and 10: mild severity*
*Score between 10 and 15: moderate severity*
*Score over 15: high severity*

## Appendix 2: Table of results according to HEADSSS questionnaire items in adolescents with no etiology found

**Appendix 2 : Table of results according to HEADSSS questionnaire items in adolescents with no etiology found**
|  | **Total**<br>**sample**<br>**size**<br>**(%)** | **Without**<br>**etiology**<br>**females**<br>**(%)** | **Without**<br>**etiology**<br>**males**<br>**(%)** | **p\*** | **Effect size**<br>**[95.0 % CI]** |
| --- | --- | --- | --- | --- | --- |
| **Single-parent**<br>**family** | 35 / 60<br>(58.3) | 23 / 36<br>(63.9) | 4 / 8<br>(50.0) | 0.742 | 0.28 [-0.49, 1.05] |
| **Siblings** | 150 / 155<br>(96.8) | 53 / 54<br>(98.1) | 16 / 16<br>(100.0) |  |  |
| **Family tensions** | 48 / 78<br>(61.5) | 33 / 49<br>(67.3) | 5 / 13<br>(38.5) | 0.114 | 0.6 [-0.02, 1.22] |
| **Resource**<br>**person** | 20 / 27<br>(74.1) | 15 / 20<br>(75.0) | 1 / 1<br>(100.0) |  |  |
| **Recent move** | 16 / 34<br>(47.1) | 12 / 19<br>(63.2) | 3 / 9<br>(33.3) | 0.283 | 0.63 [-0.18, 1.44] |
| **New person at**<br>**home** | 14 / 21<br>(66.7) | 9 / 15<br>(60.0) | 1 / 2<br>(50.0) | 1 | 0.2 [-1.28, 1.68] |
| **Known school**<br>**year and**<br>**stream** | 172 / 2234<br>(7.7) | 76 / 654<br>(11.6) | 22 / 289<br>(7.6) | 0.081 | 0.14 [0.0, 0.28] |
| **Good grades** | 19 / 22<br>(86.4) | 11 / 12<br>(91.7) | 3 / 5<br>(60.0) | 0.388 | 0.8 [-0.28, 1.88] |
| **A change in**<br>**ratings** | 5 / 7<br>(71.4) | 3 / 4<br>(75.0) | 1 / 1<br>(100.0) |  |  |
| **Plans for the**<br>**future** | 18 / 19<br>(94.7) | 13 / 13<br>(100.0) | 1 / 2<br>(50.0) | 0.264 |  |
| **School friends** | 33 / 41<br>(80.5) | 22 / 28<br>(78.6) | 6 / 7<br>(85.7) | 1 | 0.19 [-0.64, 1.02] |
| **Frequent**<br>**absenteeism** | 28 / 39<br>(71.8) | 12 / 20<br>(60.0) | 3 / 5<br>(60.0) | 1 | 0.0 [-0.98, 0.98] |
| **Like their**<br>**appearance** | 1 / 5<br>(20.0) | 0 / 4<br>(0.0) | 1 / 1<br>(100.0) | 0.401 | 0.0 [-2.19, 2.19] |
| **Recent weight**<br>**change** | 127 / 280<br>(45.4) | 31 / 77<br>(40.3%) | 12 / 39<br>(30.8) | 0.425 | 0.2 [-0.19, 0.59] |
| **Good appetite** | 242 / 709<br>(34.1) | 69 / 158<br>(43.7) | 29 / 78<br>(37.2) | 0.417 | 0.13 [-0.14, 0.4] |
| **Have hobbies** | 36 / 42<br>(85.7) | 18 / 23<br>(78.3) | 10 / 11<br>(90.9) | 0.671 | 0.36 [-0.36, 1.08] |
| **Participate in a**<br>**club activity** | 84 / 102<br>(82.4) | 30 / 43<br>(69.8) | 29 / 31<br>(93.5) | 0.026 | 0.65 [0.18, 1.12] |

**Appendix 2 (continued) : Table of results according to HEADSSS questionnaire items in adolescents with no etiology found**
|  | **Total**<br>**sample**<br>**size**<br>**(%)** | **Without**<br>**etiology**<br>**females**<br>**(%)** | **Without**<br>**etiology**<br>**males**<br>**(%)** | **p\*** | **Effect size**<br>**[95.0 % CI]** |
| --- | --- | --- | --- | --- | --- |
| **Uses**<br>**substances** | 33 / 121<br>(27.3) | 11 / 57<br>(19.3) | 9 / 22<br>(40.9) | 0.090 | 0.48 [-0.02, 0.98] |
| **In relationship** | 19 / 24<br>(79.2) | 11 / 15<br>(73.3) | 4 / 4<br>(100.0) | 0.636 |  |
| **Viergo** | 56 / 83<br>(67.5) | 30 / 47<br>(63.8) | 1 / 1<br>(100.0) |  |  |
| **Heterosexual** | 0 / 2<br>(0.0) | 0 / 1<br>(0.0) | 0 / 0 |  |  |
| **Practice sexe**<br>**safe** | 10 / 24<br>(41.7) | 4 / 12<br>(33.3) | 1 / 1<br>(100.0) | 0.805 |  |
| **Takes the pill** | 31 / 83<br>(37.3) | 16 / 50<br>(32.0) | 0 / 0 |  |  |
| **Has**<br>**experienced**<br>**sexual violence** | 15 / 29<br>(51.7) | 13 / 24<br>(54.2) | 0 / 2<br>(0.0) | 0.461 |  |
| **Sadness** | 40 / 64<br>(62.5) | 29 / 44<br>(65.9) | 5 / 11<br>(45.5) | 0.367 | 0.42 [-0.25, 1.09] |
| **Suicidal**<br>**thoughts** | 16 / 50<br>(32.0) | 11 / 37<br>(29.7) | 2 / 6<br>(33.3) | 1 | 0.08 [-0.78, 0.94] |
| **History suicide**<br>**attempt** | 32 / 48<br>(66.7) | 17 / 30<br>(56.7) | 6 / 7<br>(85.7) | 0.320 | 0.68 [-0.16, 1.52] |
| **History of**<br>**self-harm**<br>**(cutting)** | 30 / 50<br>(60.0) | 22 / 37<br>(59.5) | 5 / 8<br>(62.5) | 1 | 0.06 [-0.7, 0.82] |
| **Victim of**<br>**bullying** | 42 / 66<br>(63.6) | 25 / 40<br>(62.5) | 8 / 12<br>(66.7) | 1 | 0.09 [-0.56, 0.74] |
| **Victim of de**<br>**physical**<br>**violence** | 18 / 35<br>(51.4) | 10 / 20<br>(50.0) | 4 / 5<br>(80.0) | 0.480 | 0.66 [-0.34, 1.66] |

*p\* : With a p-value ≤ 0.05*

## Appendix 3: Results according to HEADSSS questionnaire items in the study population

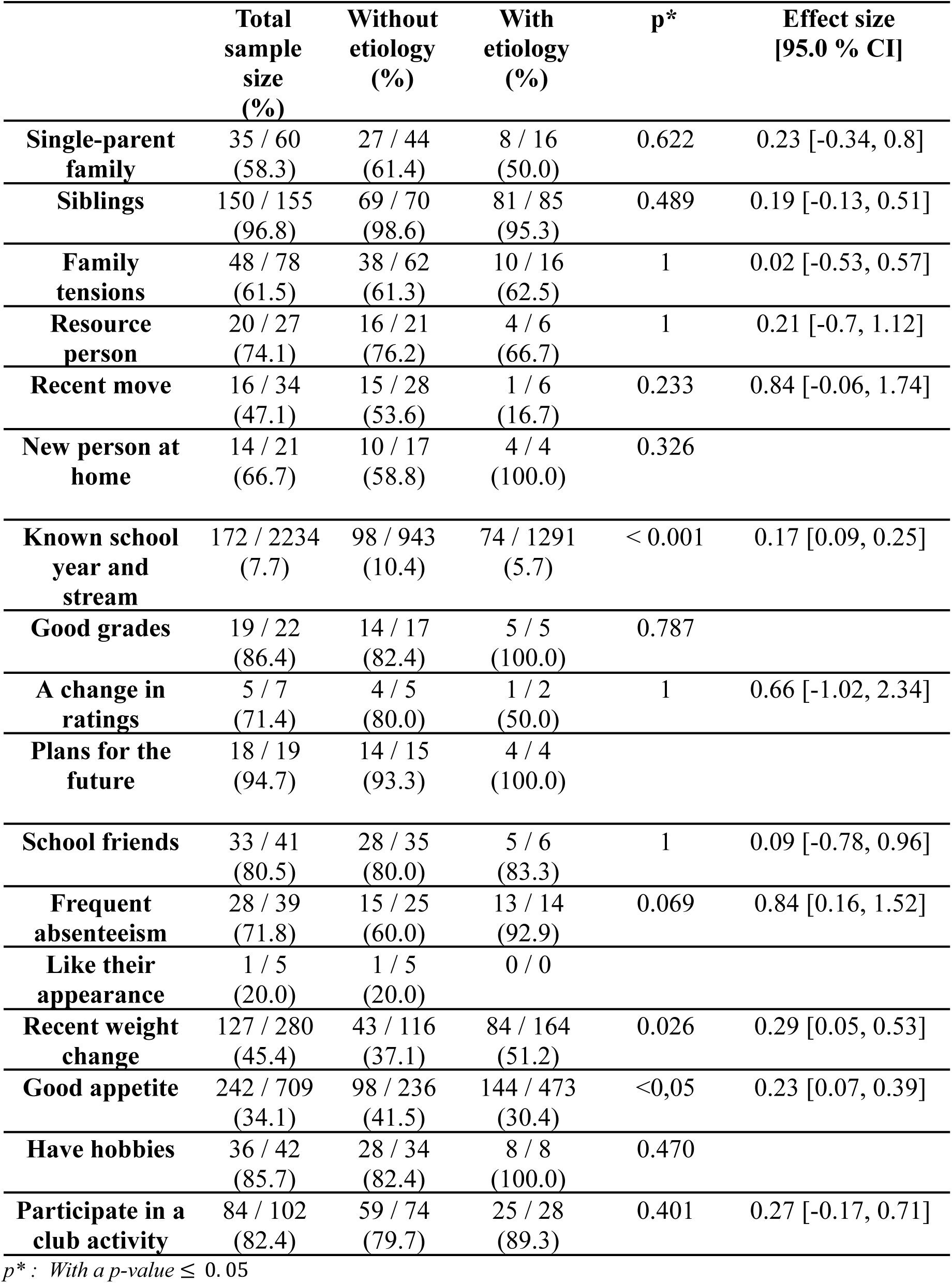
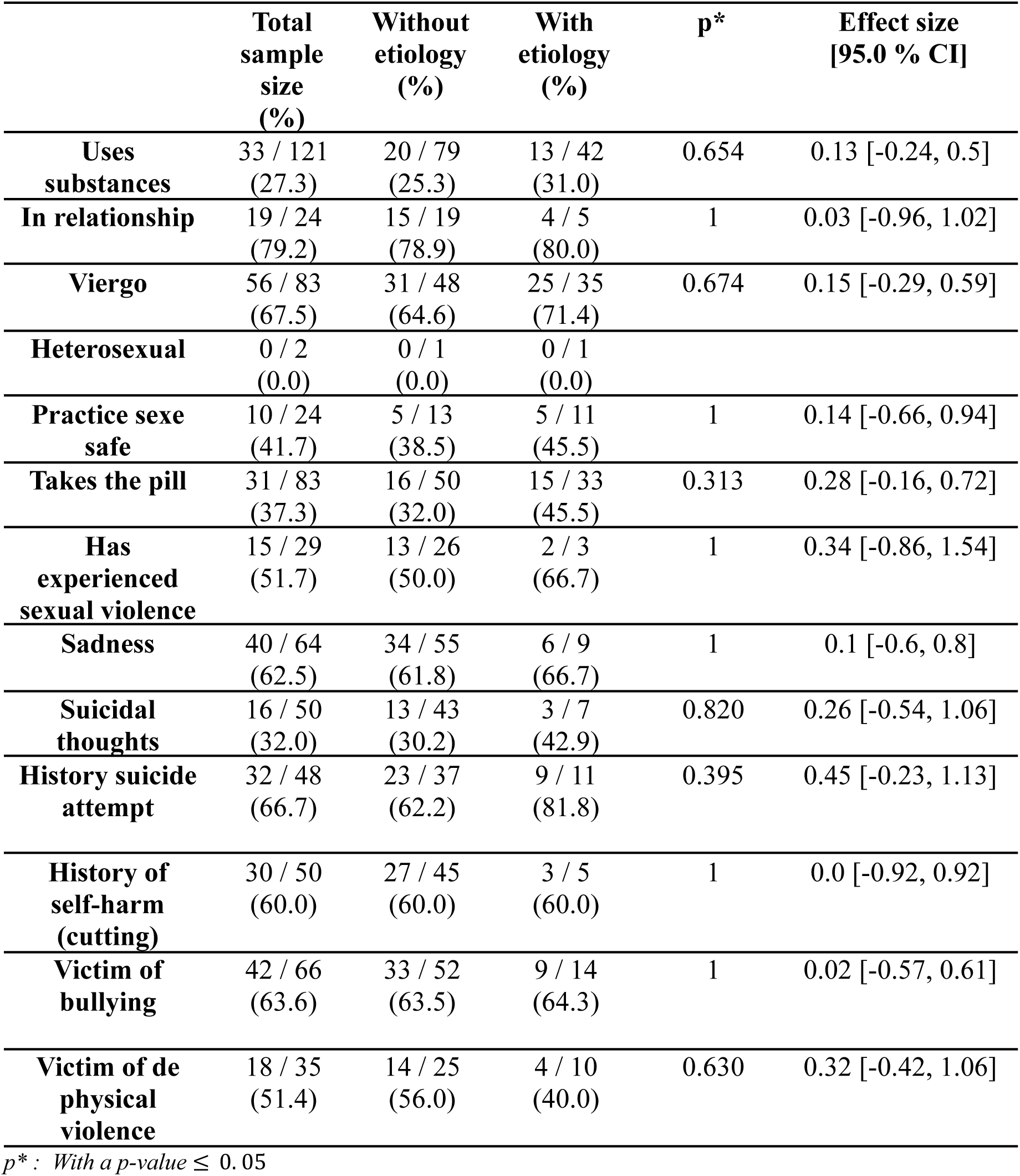

## Appendix 4: Number of adolescents by symptom with or without etiology 5A for female and 5B for boys

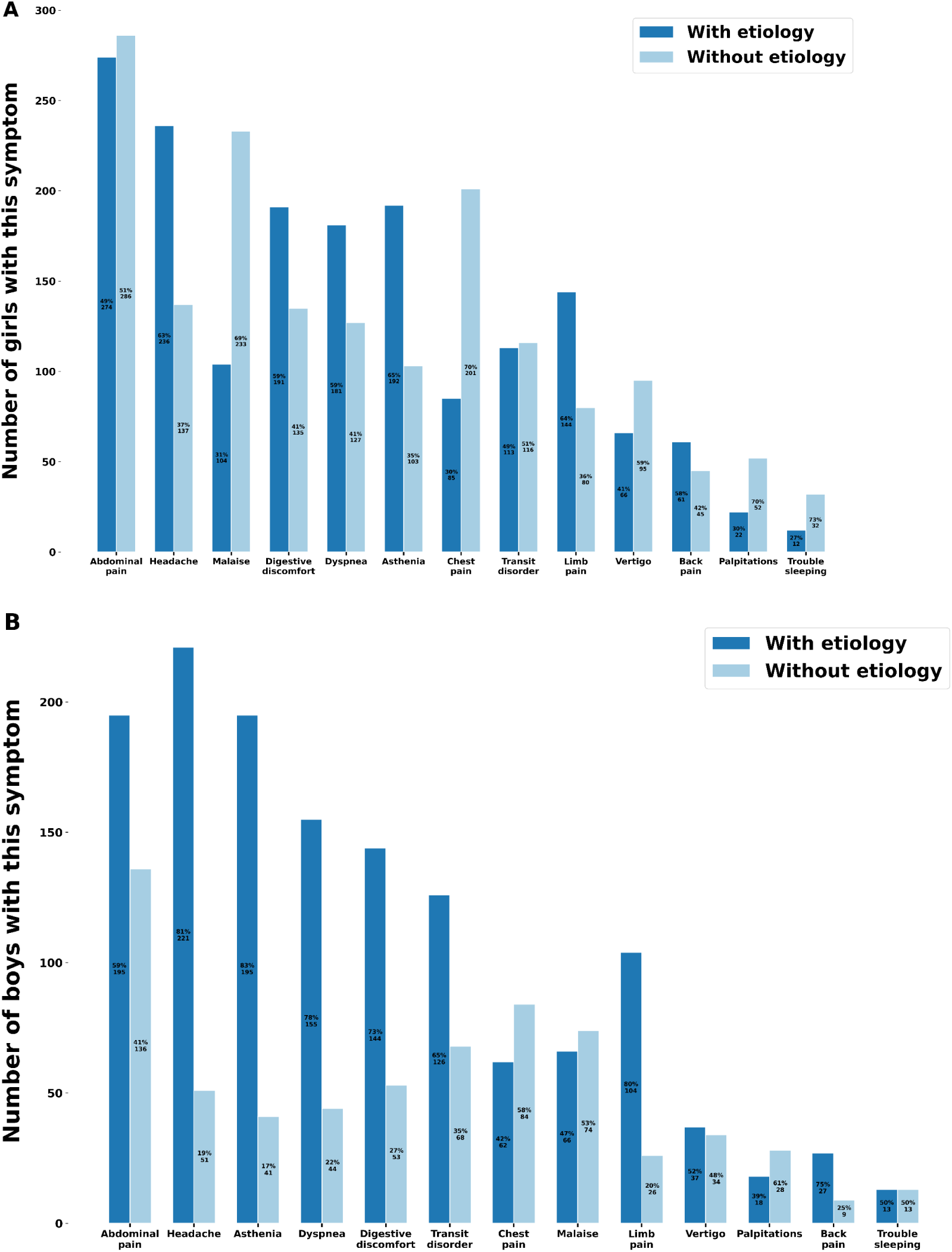

## Appendix 5: Score HEADS-ED

Appendix 5 : Score HEADS-ED
| **HEADS-ED** | **0**<br>No action needed | **1**<br>Needs action<br>but not immediate | **2**<br>Needs immediate action |
| --- | --- | --- | --- |
| **Home**<br>*Example: How does your family get along with each other?* | ○ Supportive | ○ Conflicts | ○ Chaotic / dysfunctional |
| **Education**<br>*Example: How is your school attendance? How are your grades?* | ○ On track | ○ Grades dropping / absenteeism | ○ Failing / not attending school |
| **Activities**<br>*Example: What are your relationships like with your friends?* | ○ No change | ○ Reduced / peer conflicts | ○ Fully withdrawn / significant peer conflicts |
| **Drugs & alcohol**<br>*Example: How often are you using drugs or alcohol?* | ○ None or infrequent | ○ Occasional | ○ Frequent / daily |
| **Suicidality**<br>*Example: Do you have any thoughts of wanting to kill yourself?* | ○ No thoughts | ○ Ideation | ○ Plan or gesture |
| **Emotions, behaviours, thought disturbance**<br>*Example: How have you been feeling lately?* | ○ Mildly anxious / sad / acting out | ○ Moderately anxious / sad / acting out | ○ Significantly distressed / unable to function / out of control / bizarre thoughts |
| **Discharge resources**<br>*Example: Do you have any help or are you waiting to receive help (counselling etc)?* | ○ Ongoing / well connected | ○ Some / not meeting needs | ○ None / on waitlist / non-compliant |

*A HEADS-ED score greater than or equal to eight and/or a suicide score equal to two were associated with a six-fold increase in the probability of requiring hospital care (41).*

